# Cost-effectiveness of romosozumab for severe postmenopausal osteoporosis at very high risk of fracture in Mexico

**DOI:** 10.1101/2024.02.16.24302926

**Authors:** Juan Pablo Diaz Martinez, Therese Aubry de Maraumont, Elly Natty Sánchez, Luis Miguel Camacho Cordero, Eric Yeh

**Author notes:** Corresponding author (JPDM).

## Abstract

**Introduction:** Thise study aims to assess the cost effectiveness of romosozumab versus teriparatide, both sequenced to denosumab, for the treatment of severe postmenopausal osteoporosis at very high risk of fractures in Mexican women.

**Methods:** A Markov model was used to assess the relative cost effectiveness of 1 year of romosozumab versus 2 years of teriparatide, both sequenced to denosumab for a total treatment duration of 5 years. Outcomes for a cohort of women with a mean age of 74 years, a T-score ≤ -2.5 and a previous fragility fracture were simulated over a lifetime horizon. The analysis was conducted from the perspective of the Mexican healthcare system and used a discount rate of 5% per annum. To inform relative fracture incidence, the bone mineral density (BMD) advantage of romosozumab over teriparatide was translated into relative risks of fracture, using relationships provided by a meta-regression of osteoporosis therapy trials. Outcomes were assessed in terms of lifetime costs (2023 Mexican pesos), quality-adjusted life years (QALYs) and life-years gained (LYs).

**Results:** Base case results showed that, compared with teriparatide/ denosumab, romosozumab/ denosumab reduced costs by $51,363 MXN per patient and yielded 0.03 additional QALYs and 0.01 LYs. Scenario analyses and probabilistic sensitivity analyses confirmed that results are robust to uncertainty in model assumptions and inputs.

**Conclusions:** Results show that romosozumab/ denosumab produces greater health benefits at a lower total cost than teriparatide/ denosumab.

## Introduction

Osteoporosis is a chronic condition characterized by reduced bone strength, leading to an increased susceptibility to fractures. The risk of fractures due to osteoporosis becomes more prominent with advancing age, particularly among postmenopausal women (1). These fractures have a significant economic impact globally, resulting in substantial direct and indirect costs (2). In Mexico, the direct cost of more than 75,000 fragility fractures in 2010 amounted to $256.2 million US dollars, and it was projected to grow by 41.7% by 2020 (3). The likelihood of experiencing a hip fracture over a lifetime was estimated to be 8.5%. Fragility fractures, which occur due to weakened bone strength from minimal trauma, can result in loss of independence for patients and increased burden for both patients and their caregivers (4). They are also associated with higher risks of disability, hospitalization, and mortality, which can persist for several years, especially in the case of hip fractures (5,6).

Approximately one-third of postmenopausal women will experience a fragility fracture associated with osteoporosis during their lifetime (7). Once a postmenopausal woman experiences her first fracture due to osteoporosis, her risk for subsequent fractures increases significantly, with a more than fivefold higher likelihood of suffering another fracture within one year (8–10). A recent study showed that nearly 18% of individuals experienced a second fragility fracture, and the median time to the occurrence of the second fracture was less than two years (11).

Severe or established osteoporosis is diagnosed when a previous fracture is present and bone mineral density (BMD), measured by dual-energy X-ray absorptiometry (DXA), is -2.5 standard deviations or more below the mean value of a health young adult (12). According to a recent consensus statement in Mexico, teriparatide and denosumab were recommended as treatment options for severe osteoporosis (13). Furthermore, until 2022 teriparatide was the only therapy approved for reimbursement in patients with severe postmenopausal osteoporosis at very high risk of fracture (14). Teriparatide, an anabolic hormone, is recommended for individuals with osteoporosis who have a history of previous hip or spinal fractures, are at a high risk of refracture, and have not responded to bisphosphonate treatments. However, it should be noted that teriparatide has not been proven to prevent hip fractures. Furthermore, in accordance with the Clinical Practice Guidelines in Mexico for the management of osteoporosis in postmenopausal women established by CENETEC (National Center for Excellence in Health Technology), hip fractures remain prevalent and do not exhibit a decline. These fractures are associated with significant morbidity, mortality, and financial burdens (15,16).

Romosozumab is a humanized monoclonal antibody which works by inhibiting the protein sclerostin. It has a dual effect on the bones: it increases bone formation and decreases bone resorption, unlike other bone-forming agents such as teriparatide, where both bone formation and resorption are increased. Compared to bone-forming agents or aniresoptives, romosozumab treatment duration is only one year. Romosozumab has undergone comprehensive investigation to evaluate its potential in reducing the risk of fractures, with a focus on two Phase III studies that incorporated primary and secondary fracture endpoints: the ARCH (17) and FRAME (18) studies. Additionally, a separate study assessed the percentage change in a real bone mineral density (BMD) at the total hip as an outcome measure (19) (STRUCTURE trial). Romosozumab has been shown to increase trabecular and cortical bone mass, improve bone structure and strength, and reduce the incidence of new vertebral fractures by 73% compared to placebo in just one year of treatment. In comparison to teriparatide, romosozumab has demonstrated to double the BMD at key sites: +4.4% in the spine, +2.4% in the hip, and +3% in the femur.

Economic modeling is frequently employed to assess the relative cost-effectiveness of pharmacological treatments for osteoporosis (20–22). Romosozumab has been subject to evaluation in various contexts within these studies (23–25). The findings of these assessments indicate that romosozumab offers a cost-effective alternative compared to teriparatide and alendronate as a first-line treatment option for postmenopausal women. However, the specific evidence regarding the cost-effectiveness of romosozumab in the Mexican healthcare system is currently lacking.

As a result, the objective of this study was to analyze the cost-effectiveness of romosozumab, in comparison with teriparatide, both administered sequentially with denosumab (romosozumab/denosumab versus teriparatide/denosumab), for women with severe postmenopausal osteoporosis who face a particularly high risk of fractures within the framework of the Mexican healthcare system. In Mexico, teriparatide is recommended following the lack of success with bisphosphonates. As a result, the sequential therapy employed did not involve an antiresorptive bisphosphonate; instead, it encompassed a selective modulator of oestrogen, receptors, denosumab, available in the Mexican national basic formulary (26).

## Methods

### Patient population

The patient population modelled in this analysis consisted of a cohort of postmenopausal osteoporotic women who were at very high risk for future fracture. In line with the ARCH trial, the model included patients with a mean age of 74 years with a femoral neck BMD T-score ≤ ― 2.5 and a history of fragility fracture. Based on previous clinical evidence, it was assumed that 50% of patients had a single previous fracture, and 50% had multiple prior fractures (27).

### Time horizon, perspective, and discount rate

Given that osteoporosis is a chronic disease, we used a lifetime time horizon to capture relevant benefits and costs associated with treatment. An annual discount rate of 5% was applied to both costs and future outcomes, in accordance with the Mexican HTA guidelines (14). Alternative discount rates were considered in scenario analyses. The analysis was conducted from the perspective of the public healthcare payer in Mexico.

### Comparators

Our model compared two arms:

- In the first arm, patients received romosozumab 210 mg monthly for 12 months, and were then sequenced to denosumab, 60 mg every six months (romosozumab/denosumab).
- In the second arm, patients received teriparatide 20 μg daily for 24 months, sequenced to denosumab, 60 mg every six months (teriparatide/denosumab)

The choice of these comparators relies on current guidelines for osteoporosis management in Mexico (13). Romosozumab was administered for one year (consistent with the ARCH and FRAME trials and the product label). Teriparatide was administered for 2 years consistent with the clinical guidelines. Patients in both arms were assumed to be treated for a total of 5 years; a commonly recommended duration for pharmacological osteoporosis therapy (28).

### Model structure

A Markov state transition model with a 6-month cycle length was used to assess costs, life years (LYs), and quality-adjusted life-years (QALYs) associated with the intervention arms. The structure of the model was based on the model created by the International Osteoporosis foundation. Such a model has been used widely as a basis to provide economic analyses of osteoporosis (29–33). Markov models are especially appropriate to use when: 1) a problem involves a continuous risk over time 2) the timing of events is important 3) events happen more than once, which is the case for our decision-problem (34).

Seven Markov health states were considered (Fig1): baseline (at risk of fracture), clinical vertebral fracture, post clinical vertebral fracture, hip fracture, post hip fracture, “other” fragility fracture, and death. All patients started at the baseline state, and throughout each cycle, they were at risk of sustaining a fracture (hip, vertebral, and other fracture) or death. A year after the event, patients with a hip or vertebral fracture transitioned into the post-hip fracture or post-vertebral fracture state. On the other hand, patients with another type of fracture returned to the “baseline” state.

The structure of the model is hierarchical, and the transition of the patient between the fracture states is based on the severity of the fracture type. Regarding the highest cost and lower utility, the most severe fracture is hip fracture, followed by vertebral fracture. Therefore, once the patient has sustained a vertebral fracture, the patient cannot go back to the baseline, or to the “other” fracture state(s). Similarly, patients that have sustained a hip fracture, cannot go back to the baseline, just move to the post hip state, or die. The reason for having a hierarchical structure is due to sustaining a clinical vertebral or hip fracture affects the cost and utility for the rest of the patient’s life. To rephrase, long-term effects are incurred in hip and clinical vertebral health states (21,24). However, the hierarchical structure that the Markov model holds results in underestimating the fracture incidence. To correct this, lower hierarchy fractures were separately estimated by multiplying the number of subjects in each higher hierarchical state, and with the incidence rate of the lower hierarchy fracture type in the model population.

### Model inputs

### Fracture risk

The risk of sustaining a fracture in the model depends on three elements:

1. The risk for an individual in the general population of incurring a fracture,
2. The increased fracture risk associated with osteoporosis (the relative risk RR), and
3. A risk reduction, if any, attributed to an intervention.

The general population risk depends on age and sex. The risk of fracture relative to the general population depends on age, bone mineral density and previous fracture. Age-specific general population rates of hip and vertebral fracture were derived from Ettinger et al (35). The incidences of other osteoporotic fractures were derived from Melton et al (36). Other osteoporotic fractures include ribs, pelvis, shaft/distal humerus, proximal humerus, clavicle, scapula, sternum, distal forearm, tibia,1 and fibula fractures. For age-specific incidence, these values were linearly interpolated or extrapolated as required to produce fracture rates for each year of age. General population fracture rates used in the model are shown in Supplementary Table 1.

To estimate fracture risks in untreated patients with severe postmenopausal osteoporosis, general population fracture rates were adjusted for the lower BMD T-scores and higher prevalence of previous fracture in the modelled population. Fracture risks for the patient population were adjusted with RRs of subsequent fracture in patients with a prior vertebral fracture and RRs of subsequent fracture per standard deviation decline in BMD, as described in previous economic evaluations (24,32).

### Treatment efficacy

For individuals undergoing treatment, the model incorporated data on the effectiveness of the treatment, specifically in terms of the RRs of experiencing fractures. These RRs were applied over time to the fracture rates observed in untreated very high-risk postmenopausal osteoporotic patients. The efficacy data utilized in the model provided separate RRs for hip fractures, new vertebral fractures, and nonvertebral fractures. The RRs associated with new vertebral fractures were utilized to determine the treatment-specific efficacy in preventing vertebral fractures. Similarly, the RRs of nonvertebral fractures informed the efficacy of the treatment in preventing other types of fractures, including those affecting the wrist and all other non-hip, non-vertebral fractures. The efficacy of preventing hip fractures was directly informed by the treatment-specific RRs of hip fracture.

For patients receiving romosozumab/denosumab, relative fracture incidence compared with no treatment were derived by applying the RRs for romosozumab/denosumab versus denosumab taken from a propensity-score matching (PSM) post-hoc analysis of the FRAME trial (37) to the RRs for denosumab versus placebo from a network meta-analysis of osteoporosis therapies (38) (Table 1). To allow for changing relative efficacy over time, parametric survival functions were fitted to time-to-event data for hip and nonvertebral fractures (FRAME trial). Therefore, RRs were calculated for each 6-month model cycle over the course of the 5-year treatment period. For each arm of the FRAME trial (romosozumab/denosumab and denosumab), model fitting was applied for each fracture type (hip and nonvertebral fractures). The parametric functional form with the lowest Akaike information criterion (AIC) was selected, from exponential, Weibull, log-logistic, log-normal, Gompert, generalized gamma and gamma distributions. Selected regression parameters are shown in Supplementary Table 2. This was not possible for vertebral fractures, given that accurate time-to-event data were not available. Therefore, the RR of vertebral fractures for romosozumab/denosumab versus denosumab were calculated from new vertebral fracture incidence data from the FRAME trial, shown in Table 1. Efficacy estimates for romosozumab/denosumab versus placebo from this exercise (i.e. applying RRs romosozumab/denosumab versus denosumab to RRs for denosumab versus placebo) are shown in Table 1.

**Table 1.**
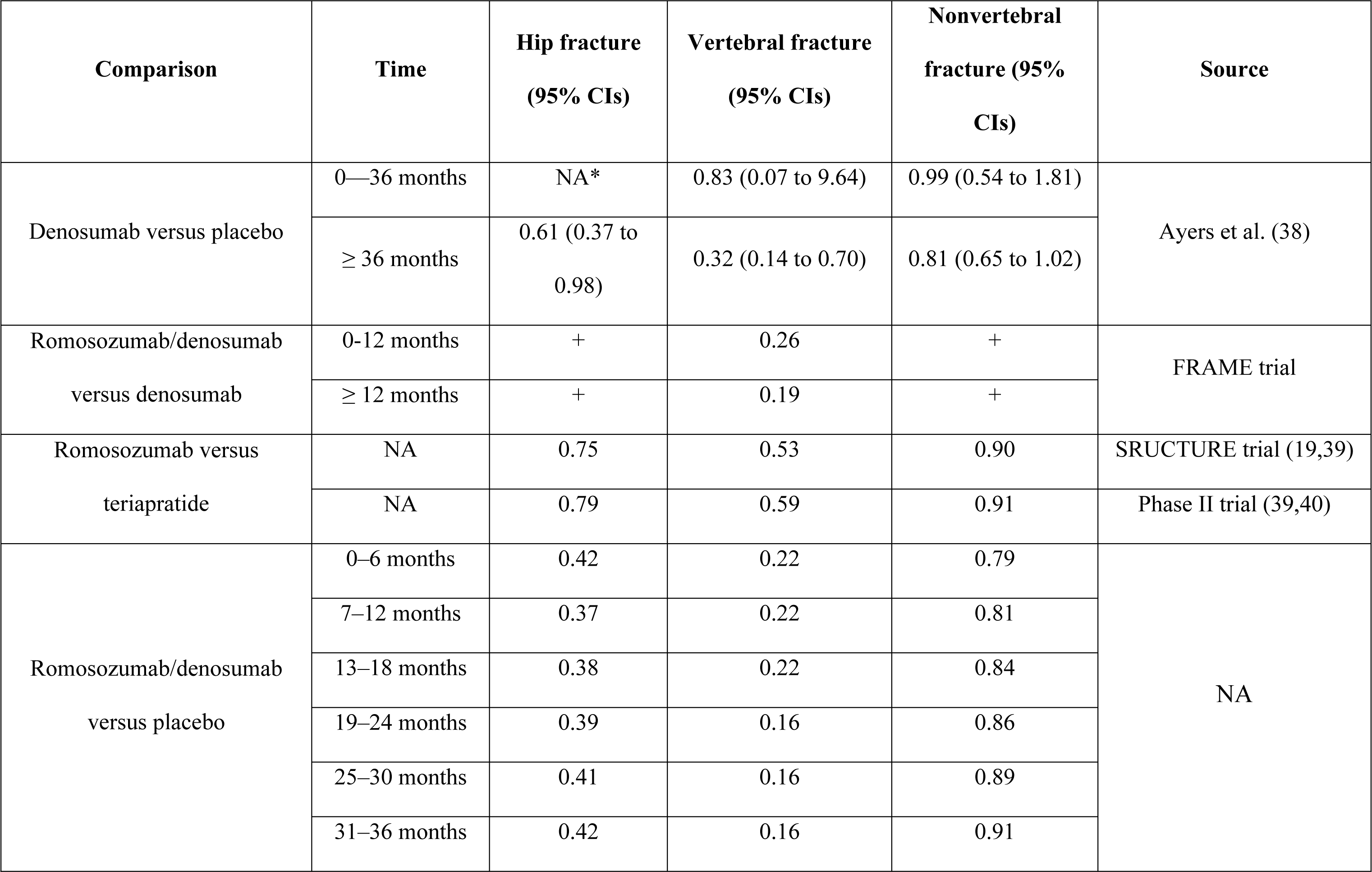
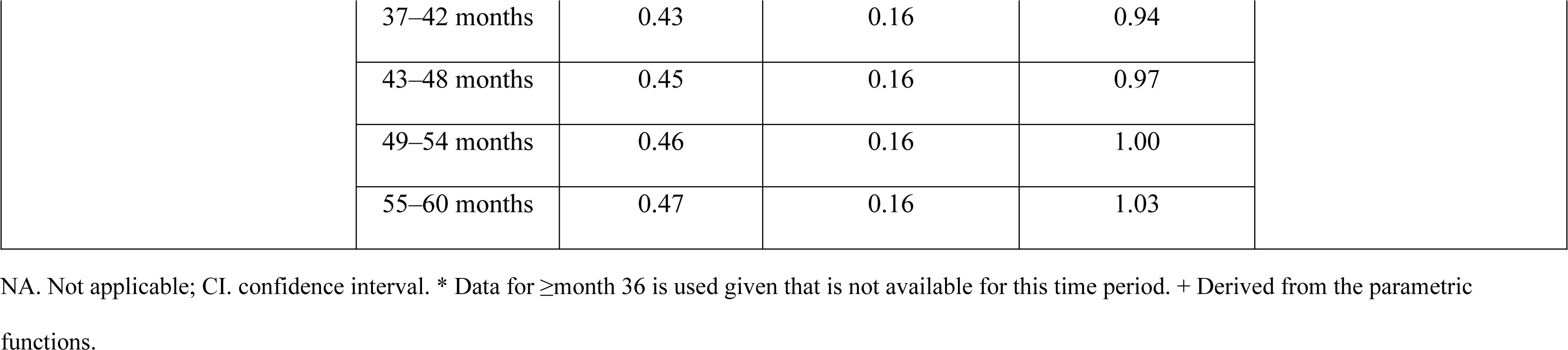
Relative risks of fractures used in the model.

| Comparison | Time | Hip fracture<br>(95% CIs) | Vertebral fracture<br>(95% CIs) | Nonvertebral<br>fracture (95%<br>CIs) | Source |
| --- | --- | --- | --- | --- | --- |
| Denosumab versus placebo | 0—36 months | NA* | 0.83 (0.07 to 9.64) | 0.99 (0.54 to 1.81) | Ayers et al. (38) |
|  | ≥ 36 months | 0.61 (0.37 to<br>0.98) | 0.32 (0.14 to 0.70) | 0.81 (0.65 to 1.02) |  |
| Romosozumab/denosumab<br>versus denosumab | 0–12 months | + | 0.26 | + | FRAME trial |
|  | ≥ 12 months | + | 0.19 | + |  |
| Romosozumab versus<br>teriapratide | NA | 0.75 | 0.53 | 0.90 | SRUCTURE trial (19,39) |
|  | NA | 0.79 | 0.59 | 0.91 | Phase II trial (39,40) |
| Romosozumab/denosumab<br>versus placebo | 0–6 months | 0.42 | 0.22 | 0.79 | NA |
|  | 7–12 months | 0.37 | 0.22 | 0.81 |  |
|  | 13–18 months | 0.38 | 0.22 | 0.84 |  |
|  | 19–24 months | 0.39 | 0.16 | 0.86 |  |
|  | 25–30 months | 0.41 | 0.16 | 0.89 |  |
|  | 31–36 months | 0.42 | 0.16 | 0.91 |  |

|  |  |  |  |  |
| --- | --- | --- | --- | --- |
|  | 37–42 months | 0.43 | 0.16 | 0.94 |
|  | 43–48 months | 0.45 | 0.16 | 0.97 |
|  | 49–54 months | 0.46 | 0.16 | 1.00 |
|  | 55–60 months | 0.47 | 0.16 | 1.03 |

NA. Not applicable; CI. confidence interval. \* Data for ≥ month 36 is used given that is not available for this time period. + Derived from the parametric functions.

For patients in the teriparatide/ denosumab arm, the efficacy in reducing fractures for this arm compared to no treatment was determined by applying the RRs of fracture for teriparatide versus romosozumab to the RRs for romosozumab/ denosumab versus placebo (described in the previous paragraph).

To derive the RRs of fracture for teriparatide versus romosozumab, BMD efficacy results from the STRUCTURE trial (19) were converted to fracture outcomes using the relationship between percentage total hip BMD change from baseline and RRs of hip, vertebral, and nonvertebral fracture on the log scale, provided by the meta-regression conducted by the FNIH Bone Quality Study (39). While the regression equations were not directly reported in the publication, fracture risk reductions associated with a 2%, 4%, and 6% improvement in BMD were available, which were used to reproduce the parameters. The slopes of these equations were used to translate the difference of 3.4% total hip BMD change from baseline for romosozumab versus teriparatide at 12 months in the STRUCTURE trial into RRs of fracture. BMD at the total hip was used in these calculations due to its high predictive value for fractures (39). Resulting RRs of fracture for romosozumab versus teriparatide are shown in Table 1. Full details of their derivation are shown in Supplmentary Table 3. Given that the STRUCTURE trial exclusively offers relative BMD efficacy data for romosozumab versus teriparatide at the 12-months, the model’s base case involves assuming that the fracture reduction advantage of romosozumab over teriparatide endures until patients in both arms transition to denosumab, with the assumed duration being 2 years. Following this period, the model posits equal efficacy between romosozumab/ denosumab and teriparatide/ denosumab. This assumption was explored in scenario analysis.

**Table 2.** Health-related quality of life, utilities, and drug, treatment monitoring/administration, direct medical costs.

| Input | Value | Source |
| --- | --- | --- |
| **Annual drug costs** |  |  |
| Romosozumab | \$52,248 | Amgen Mexico |
| Teriparatide | \$54,489 | (47). |
| Denosumab | \$7,187 | |
| **Treatment management costs** |  |  |
| Bone mineral density measurement | \$240 | (48) |
| Administration visit (for romosozumab and denosumab) | \$85 | (49) |
| Physician visit | \$1,088 | (48) |
| **Medical cost of fracture** |  |  |
| Hip fracture year 1 | \$130,177 | (50) |
| Hip fracture year 2+ | \$12,244 | |
| Vertebral fracture year 1 | \$72,530 | |
| Vertebral fracture 2+ | \$9,984 | |
| Other fracture | \$81,548 | |
| **Utility multipliers** |  |  |
| Hip fracture - 1st year after fracture | 0.55 | (46) |
| Hip fracture - 2nd and following years after fracture | 0.86 |  |
| Vertebral fracture - 1st year after fracture | 0.68 |  |
| Vertebral fracture - 2nd and following<br>years after fracture | 0.85 |  |
| Other fracture - 1st year after fracture | 0.83 |  |

**Table 3.**
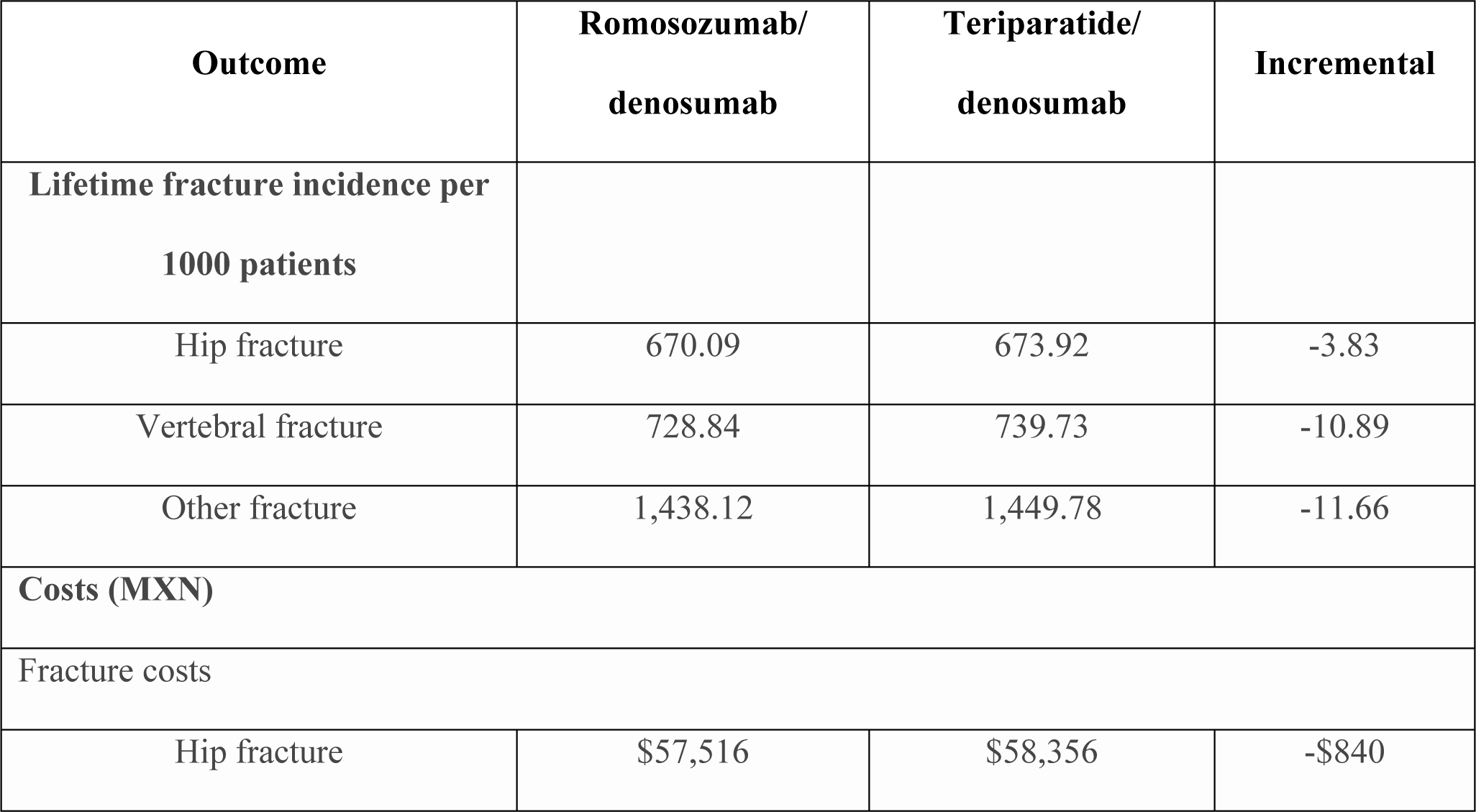
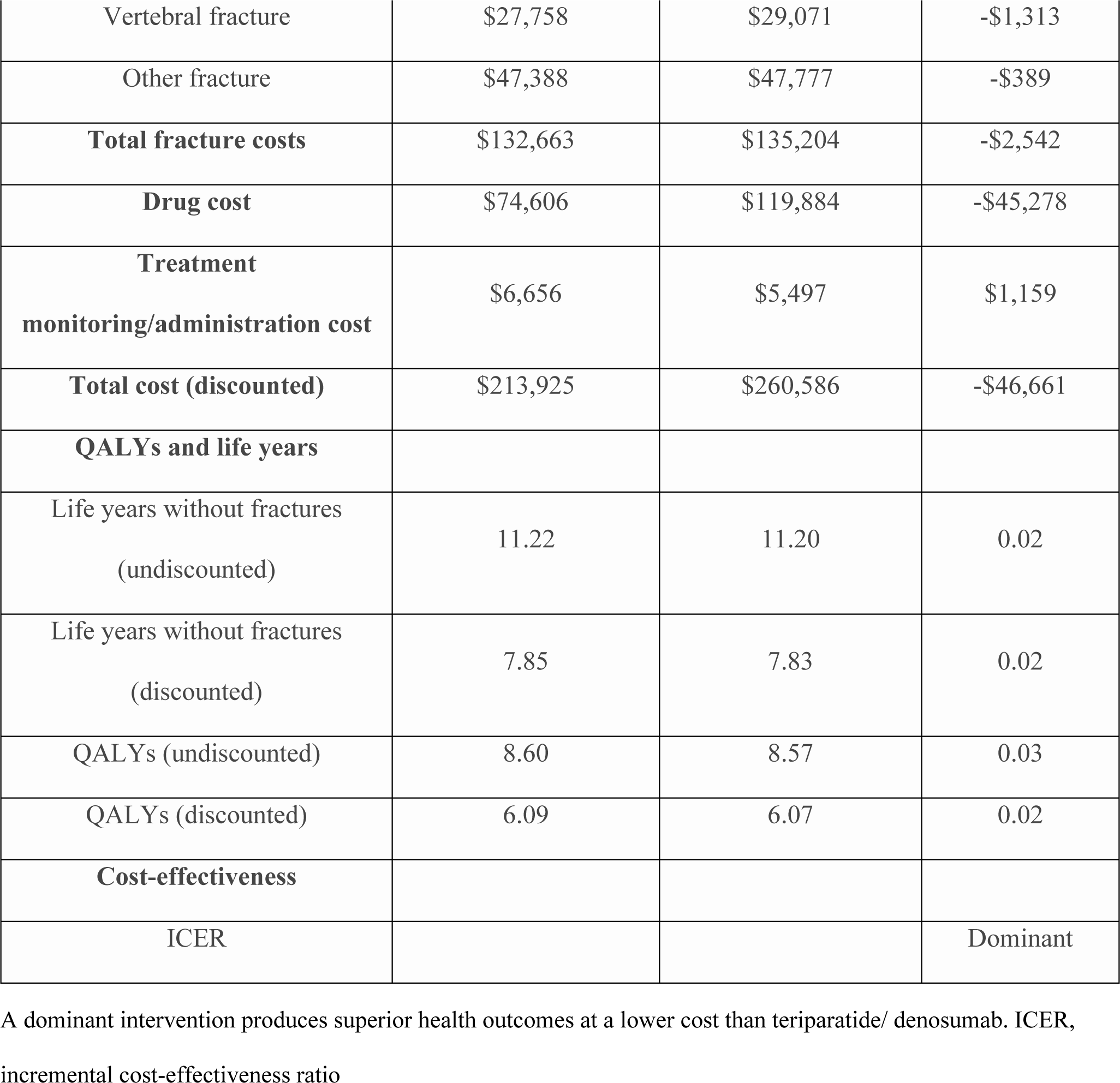
Base case lifetime cost-effectiveness results.

| Outcome | Romosozumab/<br>denosumab | Teriparatide/<br>denosumab | Incremental |
| --- | --- | --- | --- |
| **Lifetime fracture incidence per**<br>**1000 patients** |  |  |  |
| Hip fracture | 670.09 | 673.92 | -3.83 |
| Vertebral fracture | 728.84 | 739.73 | -10.89 |
| Other fracture | 1,438.12 | 1,449.78 | -11.66 |
| **Costs (MXN)** |  |  |  |
| Fracture costs |  |  |  |
| Hip fracture | \$57,516 | \$58,356 | -\$840 |
| Vertebral fracture | \$27,758 | \$29,071 | -\$1,313 |
| Other fracture | \$47,388 | \$47,777 | -\$389 |
| **Total fracture costs** | \$132,663 | \$135,204 | -\$2,542 |
| **Drug cost** | \$74,606 | \$119,884 | -\$45,278 |
| **Treatment**<br>**monitoring/administration cost** | \$6,656 | \$5,497 | \$1,159 |
| **Total cost (discounted)** | \$213,925 | \$260,586 | -\$46,661 |
| **QALYs and life years** |  |  |  |
| Life years without fractures<br>(undiscounted) | 11.22 | 11.20 | 0.02 |
| Life years without fractures<br>(discounted) | 7.85 | 7.83 | 0.02 |
| QALYs (undiscounted) | 8.60 | 8.57 | 0.03 |
| QALYs (discounted) | 6.09 | 6.07 | 0.02 |
| **Cost-effectiveness** |  |  |  |
| ICER |  |  | Dominant |

A dominant intervention produces superior health outcomes at a lower cost than teriparatide/ denosumab. ICER, incremental cost-effectiveness ratio

Finally, the STRUCTURE trial was conducted in subjects previously treated with bisphosphonates. Given that newer osteoporosis therapies are often used as secondary prevention treatments (41), this is likely to be consistent with the characteristics of patients receiving bone-forming agents in clinical practice. However, to assess the cost effectiveness of romosozumab/ denosumab versus teriparatide/ denosumab in a treatment-naïve population, a scenario analysis was conducted where BMD efficacy was taken from a romosozumab phase II trial (2.8% difference in total hip change from baseline at 12 months), conducted in patients who were not bisphosphonate pre-treated (40).

### Treatment persistence

In practice, adherence to pharmacological osteoporosis treatments is often less than perfect (42). Nevertheless, by recognizing that the controlled conditions of a randomized trial may not accurately reflect treatment discontinuation in practical settings, the model’s base case assumes full persistence with all treatments. The model then explores the impact of imperfect persistence through scenario analysis.

### Treatment offset time

Although anti-fracture efficacy is likely to persist for a period of time (offset time) after a treatment is stopped, there have been very few studies that report offset time and there is a lack of consensus on the duration of offset time (29,43). As an example, some evidence has shown that after 5 years’ treatment with alendronate followed by 5 years’ treatment with placebo, patients’ BMD remained at or above pre-treatment levels (43), indicating that the benefit of pharmacological treatment persists for some time. Therefore, our model assumes that after the 5-year treatment course, fracture reduction benefit declines linearly to 0 over the course of a further 5 years for both arms.

### Mortality

General population all-cause mortality was informed by life tables for females in Mexico from the National Institute of Statistics and Geography (INEGI) (44). The model also accounted for the increased risk of mortality following a fracture. Two key assumptions were made regarding mortality following osteoporosis-related fractures: 1) 30% of the excess mortality following a fracture was attributable to the fracture itself, in line with previous analyses (21,32) and 2) the increased risk of mortality after hip and vertebral fractures was assumed to last for 8 years as per previous analyses. This duration of excess mortality only applied to hip and vertebral fractures as other fractures were assumed to only have effects in the first year of fracture. Due to lack of Mexico-specific data related to mortality due to hip, clinical vertebral and other fractures, the default values were derived using data from Sweden (20).

### Health-related quality of life

The estimates of utility in the general population were obtained from the US (45). Utility multipliers specific to the type of fracture were applied to this background age- and sex-specific utility value. To account for the health-related quality of life (HRQoL) loss due to fracture, in the first year after hip, vertebral, and other fracture, and for the second and subsequent years after hip and vertebral fracture, utility multipliers were applied to utilities of the general population. Data specific to subsequent “other” fractures were not available. Therefore, the utility multiplier in the first year after “other” fracture was assumed to correspond to that of a distal forearm fracture. These values were taken from Svedbom et al. (46). HRQoL inputs used in the model are shown in Table 2.

### Costs

The model included drug acquisition costs, treatment monitoring/administration costs, and direct medical costs due to fracture (Table 2). All costs were expressed in 2023 Mexican pesos (MXN), inflated to 2023 values where required using the Consumer Price Index (CPI) for Mexico (51). Drug acquisition costs were obtained from the manufacturer for romosozumab and from the list prices from the Governmental Electronic System for Purchases (Compranet) (47).

For treatment monitoring/administration costs, the model assumed that patients receiving treatment incurred the cost of a physician visit once a year, and the cost of a BMD measurement every 2 years, as per on the IMSS inpatient and outpatient unit costs (48). The model assumed that all patients treated with romosozumab required a monthly outpatient visit for subcutaneous injection administration (outpatient consultation visit). This cost was obtained by the “Hospital General Gea González” inpatient and outpatient unit costs (49). Fracture morbidity costs included the direct medical cost of fracture (50), applied for the year in which fracture occur.

### Analysis

The model estimated total discounted lifetime costs, LYs (total and without fractures) and QALYs for each intervention, with cost-effectiveness assessed in terms of incremental cost-effectiveness ratios (ICERs).

Our base case analysis considered LYs without fractures gained as the effectiveness of the decision-making process. In Mexico, the willingness-to-pay threshold values are typically set at 1 to 3 times the Gross Domestic Product (GDP) per capita, which translates to approximately $200,000 to $600,000 Mexican Pesos per LY gained. Our results were assessed through a probabilistic model with 1,000 simulations, where parameters were varied simultaneously according to distributions representing their uncertainty (52). Cost-effectiveness acceptability curves (CEACs) were derived to summarize the proportion of probabilistic iterations in which each comparator was cost-effective across a range of willingness to pay per QALY-gained thresholds. In addition, sensitivity analyses using the deterministic model were performed to assess the sensitivity of results to changes in individual parameters. Parameters were varied using published confidence intervals or standard errors, where available, and by 20% above and below point estimates where measures of uncertainty were unavailable. Cost-effectiveness was assessed with the incremental net monetary benefit (INMB) for each deterministic sensitivity analysis.

Scenario analyses were conducted to assess the impact of using alternative model assumptions. Scenario analyses around modelling assumptions were considered to account for annual discount rates of 0 or 3%, treatment efficacy rates estimated from parametric models, modified treatment offset time (1 year), increased excess mortality duration, using QALYs as instead of LYs, using a different time horizon (5 years), and reduced duration of fracture reduction benefits associated with romosozumab/ denosumab. We also considered relaxing the assumption of treatment persistence, i.e., treatment discontinuation. Persistence data for denosumab were taken from an analysis of osteoporotic patients in US Medicare Fee-for-Service data reported by Singer et al. (53). Persistence with romosozumab and teriparatide were reported by Chien et al. (54).Cost-effectiveness was estimated across each intervention for each scenario.

## Results

### Base case results

The total and disaggregated base case model results are presented in Table 3. Romosozumab/ denosumab yielded 7.85 LYs without fractures, and a total cost of $213,925 MXN. Compared to teriparatide/ denosumab, it was associated with a lifetime gain of .02 LYs, and a cost reduction of $46,61 MXN. Except for treatment monitory costs, all other costs (drug and morbidity costs) were higher for those patients treated with teriparatide/ denosumab. Therefore, the improvements in QALYs and cost reductions for romosozumab/ denosumab were driven by a reduction of the expected number of fractures (2542.3 per 1000 patients versus 2572.2 for teriparatide/ denosumab). When considering QALYs, romosozumab/ denosumab mean estimate was 6.09 and teriparatide/ denosumab 6.07. Consequently, teriparatide/ denosumab was dominated by romosozumab/ denosumab independently of the measure of effectiveness.

In our probabilistic sensitivity analyses, 97% our simulations yield lower costs and more LYs without fractures for romosozumab/ denosumab when compared to teriparatide/ denosumab (Figure 2). The probability of romosozumab/ denosumab being cost-effective was 100%, independently of the threshold chosen (Figure 3).

**Figure 1.**
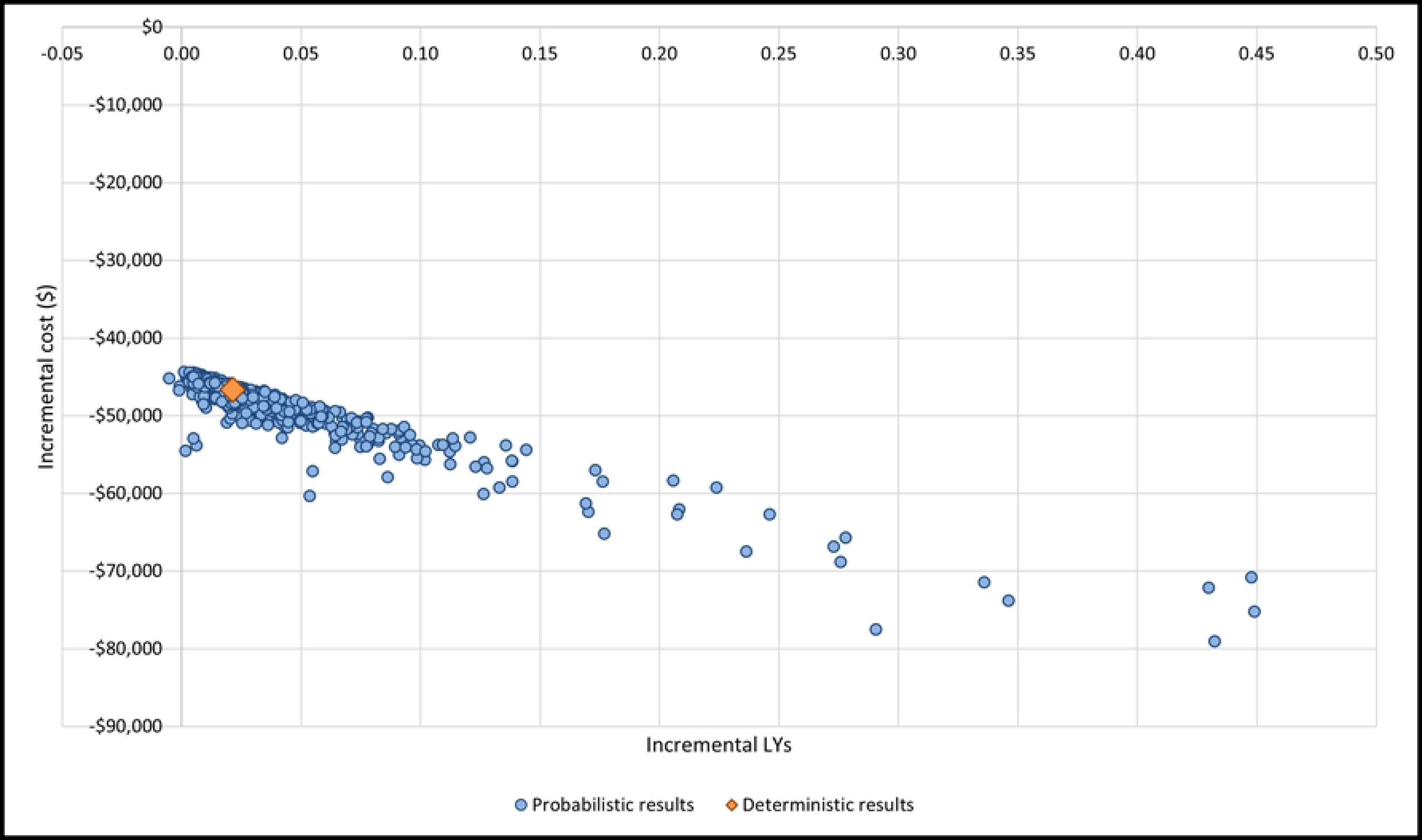
Structure of the Markov cohort model. Arrows to the health state “dead” excluded for simplification. Fx, fracture; Vert, vertebral. Death can occur from any other state. For simplicity, specific transitions are not shown.

**Figure 2.**
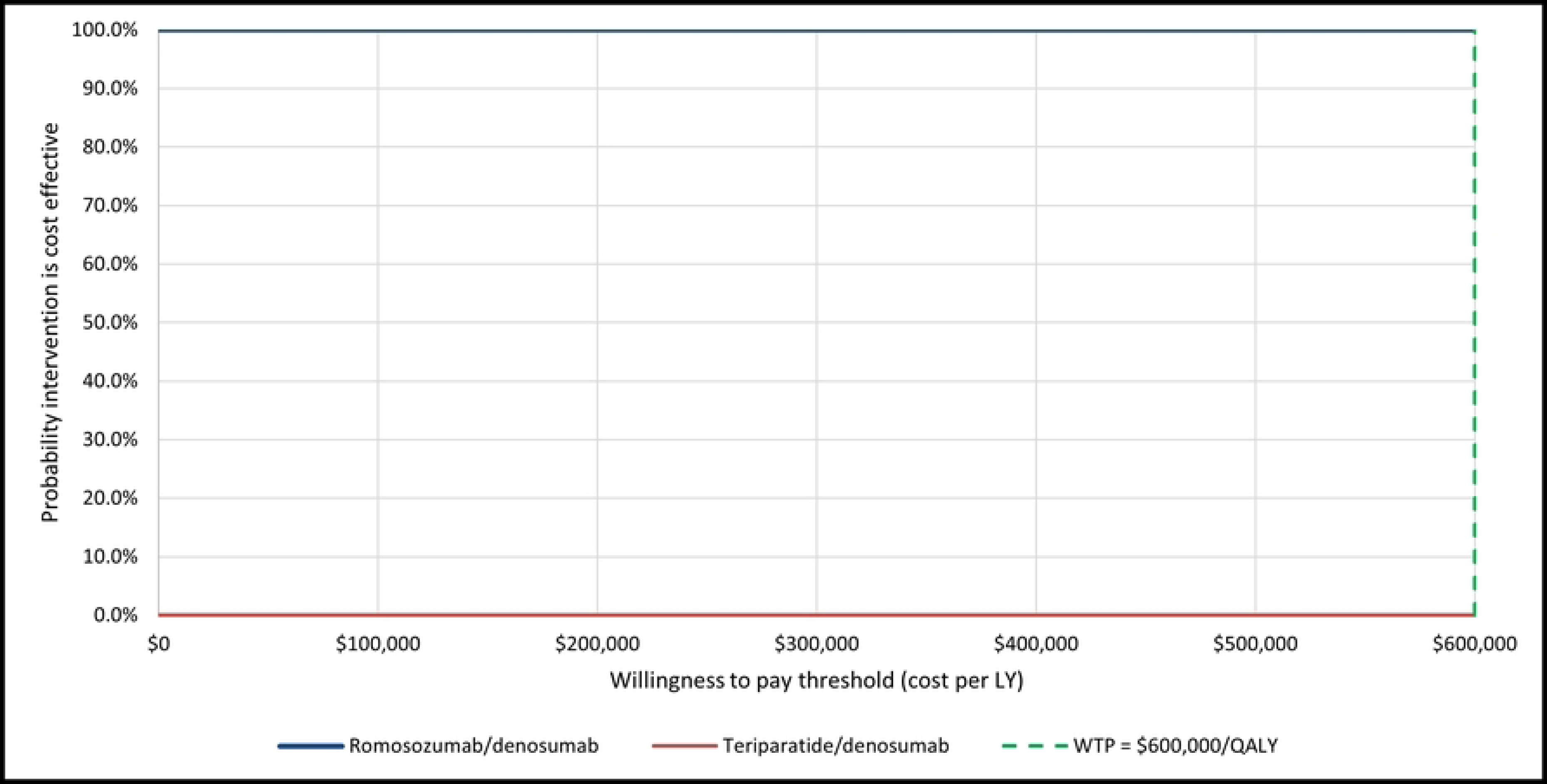
Scatter plot of probabilistic results (incremental). LY, life year

**Figure 3.**
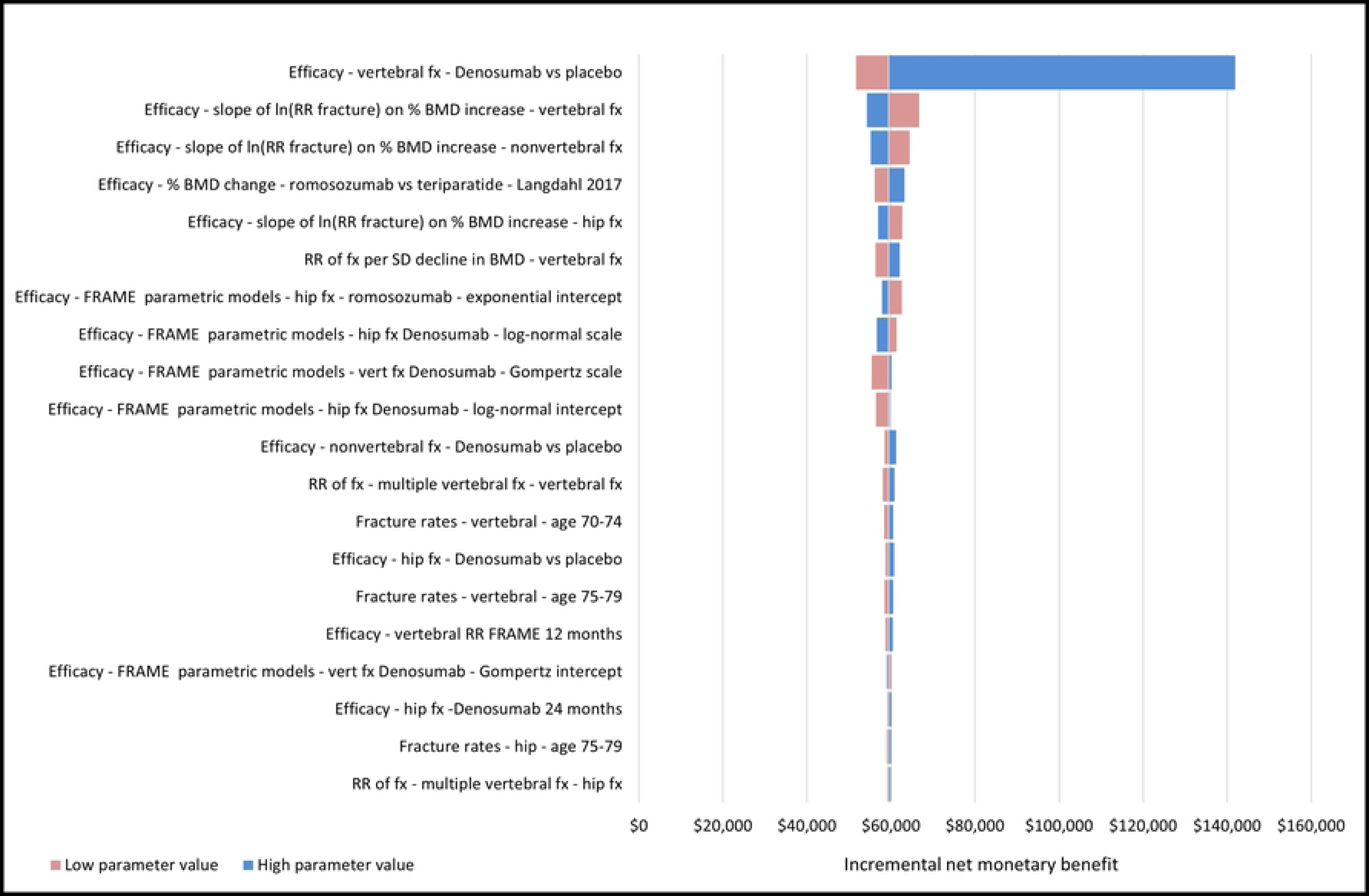
Cost-effectiveness acceptability curves. LY, life year

### Deterministic sensitivity analyses and scenario analyses

Deterministic sensitivity analysis results demonstrated that the model outcomes were the most sensitive to the RR for vertebral fractures between denosumab vs placebo. None of the cost variations proposed in this study had an impact on the model outcomes. Moreover, romosozumab/ denosumab was consistently dominant, as it represented the highest INMB relative to teriparatide/ denosumab for each run of the one-way sensitivity analysis (see Figure 4).

**Figure 4.**
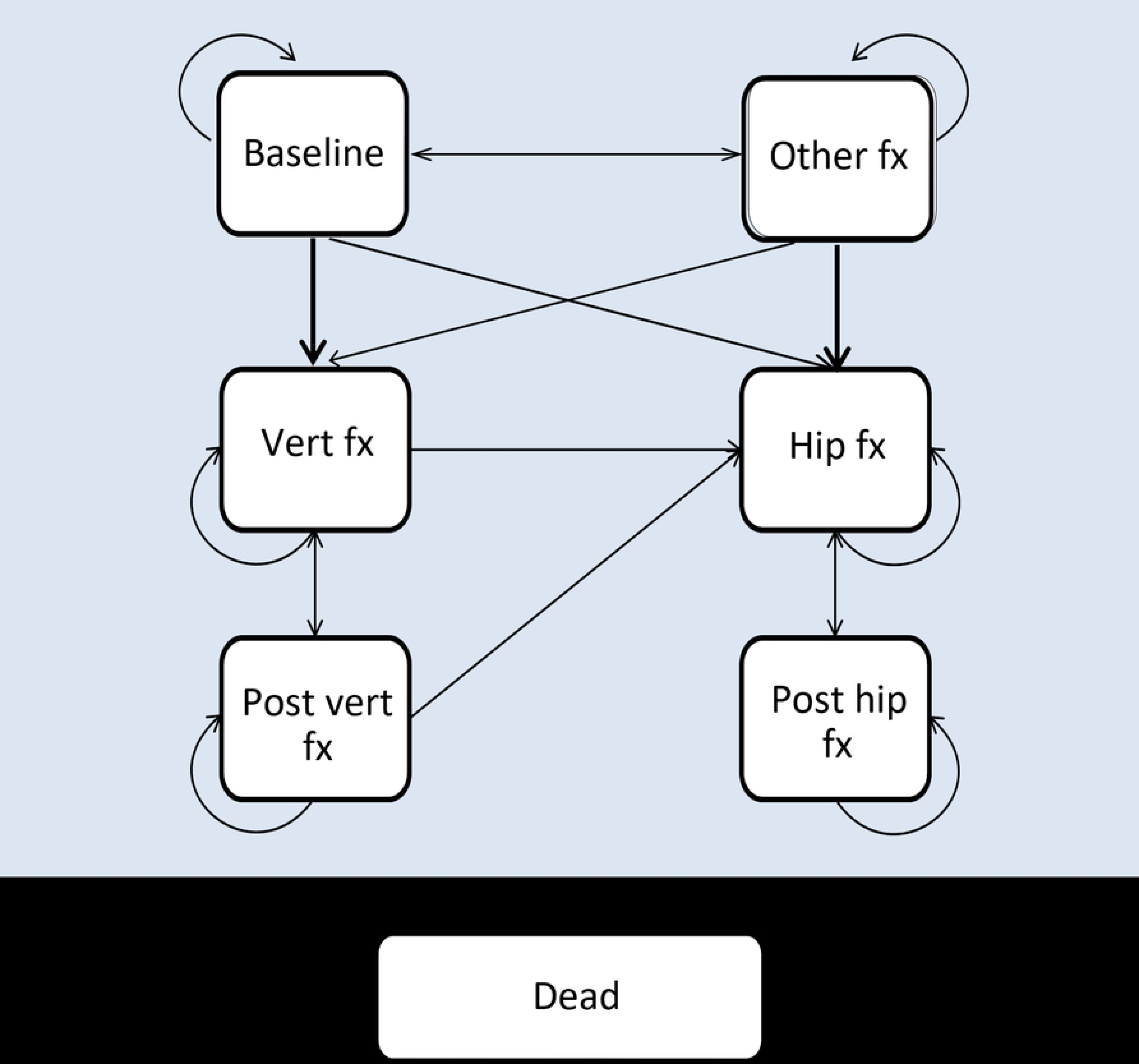
Tornado diagram of deterministic sensitivity analyses. Romosozumab/alendronate versus alendronate—discounted incremental net monetary benefit at a threshold of $600,000/QALY. FRAME, FRActure study in postmenopausal woMen with ostEoporosis; BMD, bone mineral density; Fx, fracture; QALY, quality-adjusted life year; RR, relative risk; SD, standard deviation; vert, vertebral

The incremental costs, LYs, and ICER of romosozumab/ denosumab relative to teriparatide/ denosumab for all scenarios are presented in Table 4. All scenarios yielded similar results to the base case, where romosozumab/ denosumab yielded additional LYs without fractures and fewer costs relative to teriparatide/ denosumab (i.e., remained the dominant intervention).

**Table 4.** Scenario analysis results.

| Scenario | Δ Costs | Δ LYs | ICER |
| --- | --- | --- | --- |
| Base case | -\$46,668 | 0.02 | Dominant |
| Discount rate of 0% per annum for costs and health outcomes | -\$49,755 | 0.02 | Dominant |
| Discount rate of 3% per annum for costs and health outcomes | -\$47,853 | 0.02 | Dominant |
| Time horizon set to 5 years | -\$46,254 | 0.02 | Dominant |
| Treatment offset time of 1 year | -\$46,633 | 0.02 | Dominant |
| Duration of excess mortality following hip and vertebral fracture set to 5 years | -\$46,686 | 0.02 | Dominant |
| Duration of excess mortality following hip and vertebral fracture set to 10 years | -\$46,663 | 0.02 | Dominant |
| Treatment discontinuation | -\$13,461 | 0.05 | Dominant |
| Proportion of excess mortality due to fracture event set to 10% | -\$46,863 | 0.02 | Dominant |
| Proportion of excess mortality due to fracture event set to 50% | -\$46,483 | 0.03 | Dominant |

ICER, incremental cost-effectiveness ratio; LY, life year without fractures.

## Discussion

This economic evaluation assessed the lifetime cost-effectiveness of 1 year of romosozumab sequenced to 4 years of denosumab versus teriparatide (2 years) sequenced with denosumab (3 years), for the treatment of severe osteoporosis in postmenopausal women in Mexico with a history of osteoporotic fracture and who are at very high risk for future fracture. Our base case and in all scenarios results indicate that romosozumab dominates teriparatide; meaning that it produces a gain in LYs (and QALYs) at a lower cost. This is due to the lower drug acquisition cost of romosozumab and the superior BMD efficacy of romosozumab which, translated to fracture reduction efficacy, produces both a QALY gain and a further cost reduction from avoided fractures. Deterministic and probabilistic sensitivity analyses show that model results are robust to uncertainty in parameters. When comparing romosozumab with teriparatide, romosozumab remains cost effective at a threshold of $600,000 per LY without fractures across all deterministic sensitivity analyses, with probabilistic sensitivity analysis showing a 100% probability that romosozumab is cost effective across all assessed thresholds.

The inclusion of romosozumab within the Mexican public health institutions could contribute to reduce the clinical and economic burden associated with osteoporotic fractures. Moreover, by presenting a simple administration scheme with two injections once a month, in comparison with daily injections of teriparatide, romosozumab facilitates adherence to treatment for osteoporosis, and confers more comfort to the patient.

Our cost-effectiveness analysis model is equivalent of the one that was submitted to the Canadian Agency for Drugs and Technologies in Health (CADTH) (55), and the General Health Council (Consejo de Salubridad General [CSG]) for reimbursement purposes. Although the cost effectiveness of romosozumab has already been estimated in different countries, our findings contribute to Mexico’s pharmacoeconomic literature of osteoporosis. To our knowledge, there has been only one study found that compared romosozumab with teriparatide (24). However, instead of being sequenced with denosumab, Hagino et al study used alendronate agent as the subsequent treatment (24). This specific study was conducted in Japan, and included women with severe postmenopausal osteoporosis that was treated with bisphosphonates as candidates. Hagino et al study found that romosozumab/ alendronate produced greater health benefits at a lower economic cost than teriparatide/ alendronate, which is consistent with our findings.

### Limitations and future work

As with all analyses based on economic models, this evaluation has several limitations. The most important one is that this model uses a surrogate outcome – BMD – to estimate relative fracture incidence for romosozumab versus teriparatide, rather than directly observed fracture outcomes. However, as has been demonstrated by the FNIH meta-regression, treatment-induced changes in BMD are a strong predictor of fracture outcomes. Moreover, this method is preferable to estimating relative fracture outcomes via indirect methods which, as previously discussed, would necessitate synthesis of heterogeneous trials, resulting in a high degree of uncertainty.

A further limitation is that direct BMD efficacy data for romosozumab versus teriparatide are only available for up to 12 months in the STRUCTURE and romosozumab phase II trials. This is pertinent because romosozumab and teriparatide are provided for different durations (maximum of 1 year and 2 years, respectively) prior to sequencing to denosumab, meaning there is uncertainty in relative efficacy over time. Trial-based evidence also shows that teriparatide continues to produce an improvement in total hip BMD after 12 months of treatment (57). However, even under the conservative assumption that both regimens are equally efficacious after 12 months, romosozumab/ denosumab produces equivalent health outcomes at a lower cost than teriparatide/ denosumab.

The hierarchical nature of the model structure is also a limitation. Because patients cannot experience less severe fractures once they have had a more severe fracture, the model most likely underestimates the number of fractures in the population over their lifetime. Additionally, there is uncertainty in the duration of treatment “offset time” (the duration of fracture reduction benefit after treatment discontinuation). While the duration of the offset time was based on clinical evidence (43), it is not possible to precisely quantify the duration of the offset time. This assumption was tested through scenario analyses.

Another important limitation is that international data was used where appropriate local data were not available. These included HRQoL, the baseline age of the population, and RRs used to adjust general population fracture rates for fracture history and BMD. Mexican HTA guideline specifies to perform cost-effectiveness analysis as opposed to cost-utility analysis given the lack of HRQoL studies done in the Mexican population. Nevertheless, when using LYs instead of QALYs, romosozumab was still a dominant option.

Despite the limitations, this analysis provides clear evidence of the value-for-money of romosozumab sequenced to denosumab versus teriparatide/ denosumab, with results that are robust to alternative deterministic and scenario analyses. These results would not have been possible without the approach of converting BMD efficacy to RRs of fracture, considering the inherent uncertainty associated with indirect comparisons of fracture outcomes. Results of this evaluation are specific to the Mexican healthcare system perspective. However, it is reasonable to expect romosozumab to be cost effective in any setting where the cost per course of romosozumab/ denosumab is lower than that of teriparatide/ denosumab, since the BMD advantage of romosozumab over teriparatide guarantees a QALY gain and lower total cost. More research is needed to facilitate future cost-effectiveness analyses of bone-forming agents sequenced antiresorptives. Lastly, further research on the effectiveness of treatment in other populations (i.e., male osteoporosis, patients with secondary forms of osteoporosis) could inform the potential to extrapolate the results of this analysis.

## Conclusion

This is the first economic model that evaluates the cost effectiveness of romosozumab/ denosumab as treatment for patients with postmenopausal osteoporosis from a Mexican perspective. The results from this cost-effectiveness analysis indicates that the strategy of using romosozumab for 1 year sequenced to 4 years of denosumab produces a greater number of QALYs (and LYs), reduce the number of fractures and lower the total cost when compared to 2 years of teriparatide, sequenced to 3 years of denosumab. According to probabilistic, deterministic, and scenario analysis, there is enough evidence to claim that romosozumab/ alendronate is likely to be the most cost-effective option at any decision-maker threshold. Due to this result, romosozumab should be considered as a reimbursable medication from Mexico’s public drug plan. This will aid in treating women with post-menopausal osteoporosis who has a history and is also at high risk for osteoporotic fractures.

## Transparency

### Author Contributions

TAM, EY and JPDM contributed to the design and implementation of the research, to the analysis of the results and to the writing of the manuscript; ES and LMC to the writing of the manuscript.

### Declaration of financial/other relationships

TAM, LG, EY and LMC are employed by Amgen. JPDM has served as a consultant for Amgen and has received research grants from them.

### Funding

This work was financed by Amgen, Mexico City, Mexico.

### Availability of data and materials

The datasets generated during and/or analysed during the current study are available from the corresponding author on reasonable request.

## Notes

### Funding Statement

Yes

## References

1. Tella SH, Gallagher JC. Prevention and treatment of postmenopausal osteoporosis. J Steroid Biochem Mol Biol. 2014;142:155–70.

2. Becker DJ, Kilgore ML, Morrisey MA. The societal burden of osteoporosis. Curr Rheumatol Rep. 2010;12:186–91.

3. Carlos F, Clark P, Galindo-Suárez RM, Chico-Barba LG. Health care costs of osteopenia, osteoporosis, and fragility fractures in Mexico. Arch Osteoporos. 2013;8:1–9.

4. Cooper C. The crippling consequences of fractures and their impact on quality of life. Am J Med. 1997;103(2, Supplement 1):S12–9.

5. Morin S, Lix L, Azimaee M, Metge C, Caetano P, Leslie W. Mortality rates after incident non-traumatic fractures in older men and women. Osteoporos Int. 2011;22:2439–48.

6. Brown JP, Adachi JD, Schemitsch E, Tarride JE, Brown V, Bell A, et al. Mortality in older adults following a fragility fracture: real-world retrospective matched-cohort study in Ontario. BMC Musculoskelet Disord. 2021;22:1–11.

7. Svedbom A, Hernlund E, Ivergård M, Compston J, Cooper C, Stenmark J, et al. Osteoporosis in the European Union: a compendium of country-specific reports. Arch Osteoporos. 2013;8:1–218.

8. van Geel TA, van Helden S, Geusens PP, Winkens B, Dinant GJ. Clinical subsequent fractures cluster in time after first fractures. Ann Rheum Dis. 2009;68(1):99–102.

9. Pinedo-Villanueva R, Charokopou M, Toth E, Donnelly K, Cooper C, Prieto-Alhambra D, et al. Imminent fracture risk assessments in the UK FLS setting: implications and challenges. Vol. 14, Archives of osteoporosis. Springer; 2019. p. 1–5.

10. Balasubramanian A, Zhang J, Chen L, Wenkert D, Daigle S, Grauer A, et al. Risk of subsequent fracture after prior fracture among older women. Osteoporos Int. 2019;30:79–92.

11. Adachi JD, Brown JP, Schemitsch E, Tarride JE, Brown V, Bell AD, et al. Fragility fracture identifies patients at imminent risk for subsequent fracture: real-world retrospective database study in Ontario, Canada. BMC Musculoskelet Disord. 2021;22(1):1–10.

12. Organization WH, others. Assessment of fracture risk and its application to screening for postmenopausal osteoporosis: report of a WHO study group [meeting held in Rome from 22 to 25 June 1992]. World Health Organization; 1994.

13. Clark P, Rivera FC, Sánchez LM, Gutiérrez CFM, Neri JLV, Vázquez SMC, et al. Severe osteoporosis: Principles for pharmacological therapy in Mexico. Reumatol Clínica Engl Ed. 2021;17(2):97–105.

14. CSG. Guía para la conducción de estudios de evaluación económica para la actualización del Compendio Nacional de Insumos para la Salud [Internet]. 2023. Available from: http://www.csg.gob.mx/descargas/pdf/priorizacion/cuadro-basico/guias/conduccion_estudios/GCEEE_Enero_2023.pdf

15. Hodsman AB, Bauer DC, Dempster DW, Dian L, Hanley DA, Harris ST, et al. Parathyroid hormone and teriparatide for the treatment of osteoporosis: a review of the evidence and suggested guidelines for its use. Endocr Rev. 2005;26(5):688–703.

16. CENETEC. Diagnóstico y tratamiento de osteoporosis en mujeres posmenopáusicas. Guía de Evidencias y Recomendaciones: Guía de Práctica Clínica [Internet]. Available from: http://www.cenetec-difusion.com/CMGPC/GPC-IMSS-673-18/ER.pdf

17. Saag KG, Petersen J, Brandi ML, Karaplis AC, Lorentzon M, Thomas T, et al. Romosozumab or alendronate for fracture prevention in women with osteoporosis. N Engl J Med. 2017;377(15):1417–27.

18. Cosman F, Crittenden DB, Adachi JD, Binkley N, Czerwinski E, Ferrari S, et al. Romosozumab treatment in postmenopausal women with osteoporosis. N Engl J Med. 2016;375(16):1532–43.

19. Langdahl BL, Libanati C, Crittenden DB, Bolognese MA, Brown JP, Daizadeh NS, et al. Romosozumab (sclerostin monoclonal antibody) versus teriparatide in postmenopausal women with osteoporosis transitioning from oral bisphosphonate therapy: a randomised, open-label, phase 3 trial. The Lancet. 2017;390(10102):1585–94.

20. Jönsson B, Ström O, Eisman JA, Papaioannou A, Siris E, Tosteson A, et al. Cost-effectiveness of denosumab for the treatment of postmenopausal osteoporosis. Osteoporos Int. 2011;22:967–82.

21. Parthan A, Kruse M, Yurgin N, Huang J, Viswanathan HN, Taylor D. Cost effectiveness of denosumab versus oral bisphosphonates for postmenopausal osteoporosis in the US. Appl Health Econ Health Policy. 2013;11:485–97.

22. Moriwaki K, Mouri M, Hagino H. Cost-effectiveness analysis of once-yearly injection of zoledronic acid for the treatment of osteoporosis in Japan. Osteoporos Int. 2017;28:1939–50.

23. Goeree R, Burke N, Jobin M, Brown JP, Lawrence D, Stollenwerk B, et al. Cost-effectiveness of romosozumab for the treatment of postmenopausal women at very high risk of fracture in Canada. Arch Osteoporos. 2022;17(1):71.

24. Hagino H, Tanaka K, Silverman S, McClung M, Gandra S, Charokopou M, et al. Cost effectiveness of romosozumab versus teriparatide for severe postmenopausal osteoporosis in Japan. Osteoporos Int. 2021;32:2011–21.

25. Söreskog E, Lindberg I, Kanis J, Åkesson K, Willems D, Lorentzon M, et al. Cost-effectiveness of romosozumab for the treatment of postmenopausal women with severe osteoporosis at high risk of fracture in Sweden. Osteoporos Int. 2021;32:585–94.

26. SEGOB. EDICIÓN 2021 del Libro de Medicamentos del Compendio Nacional de Insumos para la Salud. [Internet]. 2021. Available from: https://dof.gob.mx/nota_detalle.php?codigo=5616775&fecha=26/04/2021#gsc.tab=0

27. Black DM, Arden NK, Palermo L, Pearson J, Cummings SR, Group S of OFR. Prevalent vertebral deformities predict hip fractures and new vertebral deformities but not wrist fractures. J Bone Miner Res. 1999;14(5):821–8.

28. Qaseem A, Forciea MA, McLean RM, Denberg TD, Physicians* CGC of the AC of. Treatment of low bone density or osteoporosis to prevent fractures in men and women: a clinical practice guideline update from the American College of Physicians. Ann Intern Med. 2017;166(11):818–39.

29. Jönsson B, Christiansen C, Johnell O, Hedbrandt J. Cost-effectiveness of fracture prevention in established osteoporosis. Osteoporos Int. 1995;5:136–42.

30. Svedbom A, Hadji P, Hernlund E, Thoren R, McCloskey E, Stad R, et al. Cost-effectiveness of pharmacological fracture prevention for osteoporosis as prescribed in clinical practice in France, Germany, Italy, Spain, and the United Kingdom. Osteoporos Int. 2019;30:1745–54.

31. Zethraeus N, Borgström F, Ström O, Kanis J, Jönsson B. Cost-effectiveness of the treatment and prevention of osteoporosis—a review of the literature and a reference model. Osteoporos Int. 2007;18:9–23.

32. O’Hanlon CE, Parthan A, Kruse M, Cartier S, Stollenwerk B, Jiang Y, et al. A model for assessing the clinical and economic benefits of bone-forming agents for reducing fractures in postmenopausal women at high, near-term risk of osteoporotic fracture. Clin Ther. 2017;39(7):1276–90.

33. Parthan A, Kruse M, Agodoa I, Silverman S, Orwoll E. Denosumab: a cost-effective alternative for older men with osteoporosis from a Swedish payer perspective. Bone. 2014;59:105–13.

34. Hiligsmann M, Kanis JA, Compston J, Cooper C, Flamion B, Bergmann P, et al. Health technology assessment in osteoporosis. Calcif Tissue Int. 2013;93(1):1–14.

35. Ettinger B, Black D, Dawson-Hughes B, Pressman A, Melton L 3rd. Updated fracture incidence rates for the US version of FRAX®. Osteoporos Int. 2010;21:25–33.

36. Melton III LJ, Crowson C, O’fallon W. Fracture incidence in Olmsted County, Minnesota: comparison of urban with rural rates and changes in urban rates over time. Osteoporos Int. 1999;9:29–37.

37. Cosman F, Oates M, Betah D, Ferrari S, McClung MR. One Year of Romosozumab Followed by One Year of Denosumab Compared With Two Years of Denosumab: BMD and Fracture Results From the FRAME and FRAME Extension Studies. In: Oral presentation (#1055) at 2022 ASBMR annual meetings. Austin, TX;

38. Ayers C, Kansagara D, Lazur B, Fu R, Kwon A, Harrod C. Effectiveness and safety of treatments to prevent fractures in people with low bone mass or primary osteoporosis: a living systematic review and network meta-analysis for the American College of Physicians. Ann Intern Med. 2023;176(2):182–95.

39. Bouxsein ML, Eastell R, Lui LY, Wu LA, de Papp AE, Grauer A, et al. Change in bone density and reduction in fracture risk: a meta-regression of published trials. J Bone Miner Res. 2019;34(4):632–42.

40. McClung MR, Grauer A, Boonen S, Bolognese MA, Brown JP, Diez-Perez A, et al. Romosozumab in postmenopausal women with low bone mineral density. N Engl J Med. 2014;370(5):412–20.

41. Pavone V, Testa G, Giardina SM, Vescio A, Restivo DA, Sessa G. Pharmacological therapy of osteoporosis: a systematic current review of literature. Front Pharmacol. 2017;8:803.

42. Ideguchi H, Ohno S, Hattori H, Ishigatsubo Y. Persistence with bisphosphonate therapy including treatment courses with multiple sequential bisphosphonates in the real world. Osteoporos Int. 2007;18:1421–7.

43. Black DM, Schwartz AV, Ensrud KE, Cauley JA, Levis S, Quandt SA, et al. Effects of continuing or stopping alendronate after 5 years of treatment: the Fracture Intervention Trial Long-term Extension (FLEX): a randomized trial. Jama. 2006;296(24):2927–38.

44. Actuaries S of. Mortality and Other Rate Tables [Internet]. 2010. Available from: https://mort.soa.org/

45. Hanmer J, Lawrence WF, Anderson JP, Kaplan RM, Fryback DG. Report of nationally representative values for the noninstitutionalized US adult population for 7 health-related quality-of-life scores. Med Decis Making. 2006;26(4):391–400.

46. Svedbom A, Borgstöm F, Hernlund E, Ström O, Alekna V, Bianchi ML, et al. Quality of life for up to 18 months after low-energy hip, vertebral, and distal forearm fractures—results from the ICUROS. Osteoporos Int. 2018;29:557–66.

47. SHCP. Portal del Sistema Electrónico de Compras Gubernamentales [Internet]. 2023. Available from: https://compranet.hacienda.gob.mx/

48. Federación (DOF) DO de la. ACUERDO número ACDO.AS3.HCT.251022/299.P.DF dictado por el H. Consejo Técnico, en sesión ordinaria de 25 de octubre de 2022, relativo a la aprobación de los Costos Unitarios por Nivel de Atención Médica actualizados al año 2023. [Internet]. 2023. Available from: https://www.dof.gob.mx/nota_detalle.php?codigo=5672661&fecha=29/11/2022#gsc.tab=0

49. González HGG. Tabulador de Cuotas de Recuperación [Internet]. 2018. Available from: https://www.gob.mx/salud/hospitalgea/documentos/tabulador-de-cuotas-vigentes-2018

50. Carlos-Rivera, F, Guzmán-Caniupan, JA, Clark, P, Aubry de Maraumont, T, Soria-Suarez, N. Projected frequency and economic burden of incident fragility fractures during 2023 in Mexico. 2023.

51. Geografía (INEGI) IN de E y. Índice Nacional de Precios al Consumidor. Salud y Cuidado Personall [Internet]. 2023. Available from: https://www.inegi.org.mx/app/indicesdeprecios/Estructura.aspx?idEstructura=112001300040&T=%C3%8Dndices%20de%20Precios%20al%20Consumidor&ST=INPC%20Nacional%20(mensual)

52. Briggs A, Sculpher M, Claxton K. Decision modelling for health economic evaluation. Oup Oxford; 2006.

53. Singer A, Liu J, Yan H, Stad R, Gandra S, Yehoshua A. Treatment patterns and long-term persistence with osteoporosis therapies in women with Medicare fee-for-service (FFS) coverage. Osteoporos Int. 2021;32:2473–84.

54. Chien, HC, Arora, T, Oates, M, McDermott, M, Curtis, JR. Drug utilization patterns of anabolic agents - romosozumab and parathyroid hormone analogues - among postmenopausal women in the U.S., 2019-2021. In Vancouver, Canada; 2023.

55. CADTH. Romosozumab for the treatment of osteoporosis in postmenopausal women with a history of osteoporotic fracture and who are at very high risk for future fracture. [Internet]. 2022. Available from: https://www.cadth.ca/romosozumab

56. Vestergaard Kvist A, Faruque J, Vallejo-Yagüe E, Weiler S, Winter EM, Burden AM. Cardiovascular safety profile of romosozumab: a pharmacovigilance analysis of the US Food and Drug Administration Adverse Event Reporting System (FAERS). J Clin Med. 2021;10(8):1660.

57. Leder BZ, Tsai JN, Uihlein AV, Burnett-Bowie SAM, Zhu Y, Foley K, et al. Two years of Denosumab and teriparatide administration in postmenopausal women with osteoporosis (The DATA Extension Study): a randomized controlled trial. J Clin Endocrinol Metab. 2014;99(5):1694–700.

